# What risk do *Brucella* vaccines pose to humans? A systematic review of the scientific literature on occupational exposure

**DOI:** 10.1101/2023.03.16.23287350

**Authors:** Manuel Vives-Soto, Amparo Puerta-García, José-Luis Pereira, Esteban Rodríguez-Sánchez, Javier Solera

## Abstract

**Introduction:** Vaccination of cattle, sheep and goats with attenuated strains of *Brucella* is an effective measure for controlling brucellosis, although the vaccines pose risks to humans during the manufacturing process and throughout their distribution and administration to animals.

**Objective:** The aim of this study was to review the risk posed to humans by occupational exposure to vaccine strains and the measures that should be implemented to minimize this risk.

**Materials and methods:** This article reviewed the scientific literature indexed in PubMed up to January 31, 2023, following “the PRISMA guidelines”; special emphasis was placed on the vaccine strain used and the route of exposure.

**Results:** Twelve primary reports were found: six included the Rev-1 strain, three the S19, and four the RB-51 strain. Rev-1 seemed to be the most virulent. The most frequent type of exposure was needle injury during administration, while exposure accidents during vaccine manufacturing were exceptional. Prolonged contact with the pathogen, lack of information and a low adherence to personal protective equipment (PPE) use in the work environment were determining factors for becoming infected. Despite strict protection measures, a percentage of vaccine manufacturing workers developed a positive serology to the vaccine strain, which may have conferred immunity.

**Conclusions:** *Brucella* vaccines pose risk of contagion to humans from their production to their administration to cattle; therefore, protection measures should be extreme, and active surveillance of exposed workers should be implemented.

## INTRODUCTION

Human brucellosis is a zoonosis with worldwide distribution. Although the number of cases of brucellosis may be decreasing in the world, the continued presence of the disease in some endemic areas and the potential use of *Brucella* species as an agent of bioterrorism, make brucellosis a major public health hazard with important sanitary and economic repercussions (1).

Currently, cattle, sheep and goat vaccination remains an essential measure for the control of this zoonosis, and only live attenuated vaccines have shown efficacy in preventing infection in these ruminants.The strains currently used in most countries for the control of bovine brucellosis are *Brucella abortus* S19 and *Brucella abortus* RB51, while the strain used for small ruminants is *Brucella melitensis* Rev.1 (2). These strains are capable of establishing limited infection in cattle, mimicking the natural infection process by wild strains and thus conferring protection. However, these vaccines are not used in humans due to the high risk of developing acute brucellosis (3); they are capable of infecting humans with occupational exposure via the oral, nasal, conjunctival or genital routes, and by accidental needle inoculation (4). Whatever the route of entry, the infection can be symptomatic or asymptomatic, and localized or systemic. Serologic tests continue to be used to diagnose this infection, given the risk and difficulty involved in obtaining positive cultures (5).

At present, uncertainty remains about the real risk of these vaccines for those who manufacture or administer them. Despite advances in the protection measures that must be applied when there is a risk of contact with these live bacterial vaccines (6), accidental contagion still occurs. The aim of this study was to review the risk posed to humans by occupational exposure to vaccine strains and the effectiveness of the preventive measures being implemented to minimize this risk.

## MATERIAL AND METHODS

The guidelines of the PRISMA statement (Preferred Reported Items for Systematic Reviews and Meta-Analyses) were formally adopted in this review (6). The search was conducted on January 31, 2023, with no date or country restrictions, using the PubMed database. Next, titles, abstracts and full texts were independently analyzed by two investigators. The search terms used were: Brucella (Rev-1 OR Rev.1 OR S19 OR S.19 OR B19 OR B-19 OR RB51) AND Human[Mesh]. Once the relevant studies were selected, their references were searched. Case reports of non-occupational exposure to vaccine strains, cases of intentional inoculation and publications on exposure to non-vaccine strains were excluded from the study.

## RESULTS

The PRISMA Flow Diagram is shown in **figure 1**. Twelve articles were included from 132 records screened. Eleven of these articles were identified by manual search of the references found in the full text articles reviewed, and one of them was included in the study. **Table 1** lists the main publications of brucellosis cases acquired through exposure to vaccine strains. These publications are summarized below.

| Reference | Year | Country | Vaccine strain | Case/exposed | Diagnostic criteria | Exposure type |
| --- | --- | --- | --- | --- | --- | --- |
| Ollé-Goig (8) | 1987 | Spain | Rev-1 | 48/172 | Clinical, serological<sup>a</sup> | Manufacturing exposure |
| Blasco (9) | 1993 | Spain | Rev-1 | 2/NR | Blood culture | Vaccine administration |
| CDC (10) | 1998 | USA | RB51 | 4/32 | Clinical | Vaccine administration |
| Ashford (11) | 2004 | USA | RB51 | 19/26 | Clinical, blood culture<sup>b</sup> | Vaccine administration |
| Avdikou (12) | 2005 | Greece | Rev-1 | 41/NR | Serological | Vaccine administration |
| Wallach (13) | 2008 | Argentina | S19 | 21/30 | Clinical, serological | Manufacturing exposure |
| Gunes (14) | 2013 | Turkey | Rev-1 | 10/46 | Serosurveillance<sup>c</sup> | Vaccine administration |
| Proch (15) | 2018 | India | S19 | 5/12 | Serological | Vaccine administration |
| Hatcher (16) | 2018 | USA | RB51 | 1/NR | Clinical | Vaccine administration |
| Arapovic (17) | 2020 | Bosnia<br>Herzegovina | Rev-1 | 1/NR | Blood Culture<sup>d</sup> | Vaccine administration |
| Pereira (18) | 2021 | Brasil | S19 / RB51 | 7/108 | Clinical<sup>e</sup> | Vaccine administration |
| Vives-Soto (19) | 2022 | Spain | Rev-1 / S19<sup>f</sup> | 47/115 | Serosurveillance<sup>g</sup> | Manufacturing exposure |

NR = not reported. <sup>a</sup> Positive culture in 1 individual with B19 infection and in 11 individuals with Rev.1 infection. <sup>a</sup> Two positive cultures. <sup>b</sup> One positive culture taken from the inoculation site. <sup>c</sup> Two clinical infections. <sup>d</sup> Confirmed by molecular methods (PCR). <sup>e</sup> Reported through an online questionnaire. <sup>f</sup> S19 vaccine manufacturing was marginal. <sup>g</sup> No clinical infections.

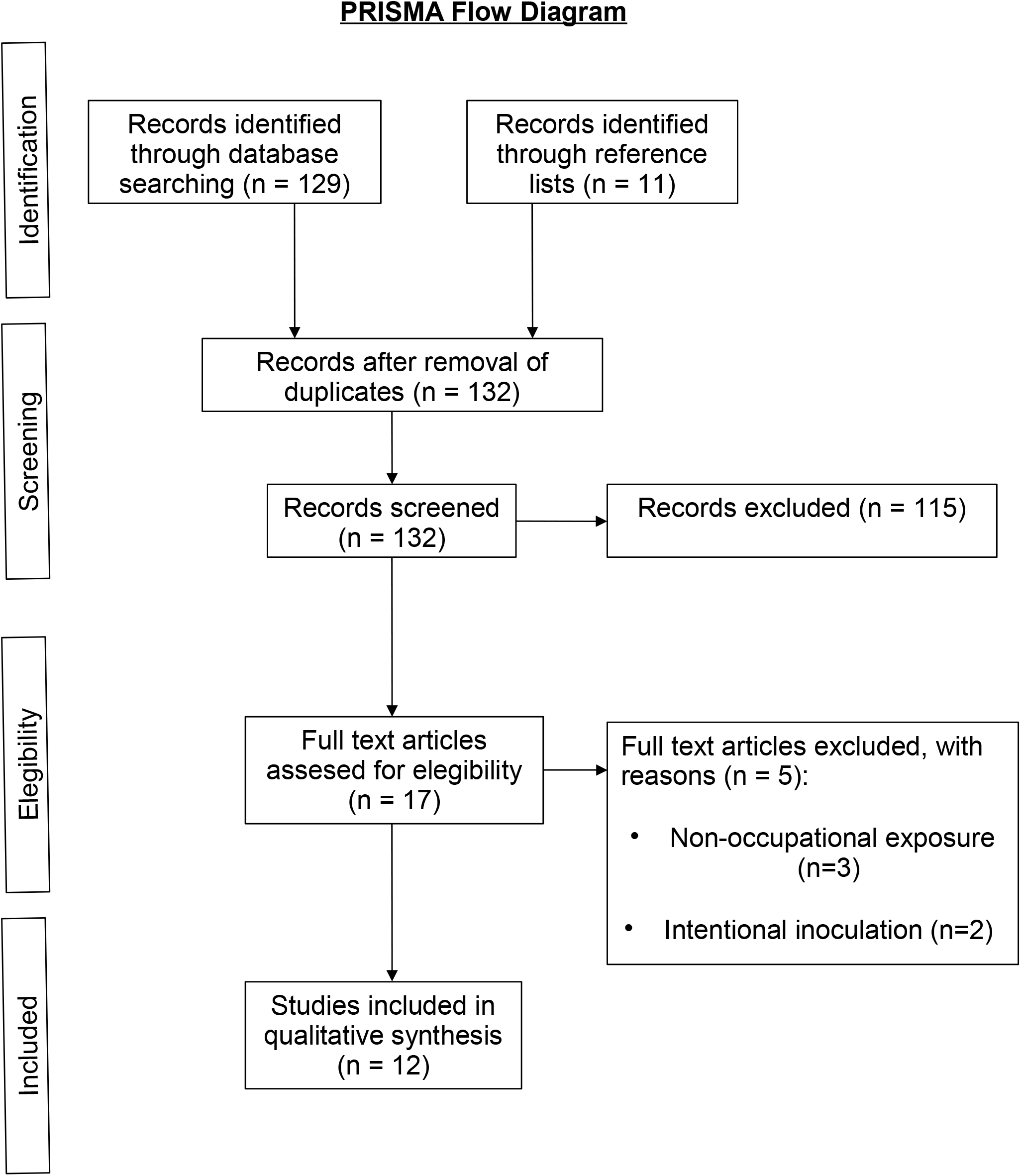

In 1987, Ollé-Goig et al. described an airborne-acquired outbreak of brucellosis in workers accidentally exposed to the Rev-1 strain at a manufacturing plant for veterinary biologic products in Gerona (Spain) (8). The study included 172 workers, of which 28 were identified as having acute brucellosis, 20 had chronic brucellosis, 106 were infection-free and 18 had no clear diagnosis. The cause of infection was a failure in the ventilation system, which resulted in an outbreak of brucellosis in nearby workers. In 1993, Blasco et al published two cases of *Brucella* infection in Spanish veterinarians accidentally exposed to the Rev-1 strain (9). In both individuals, the Rev-1 *Brucella mellitensis* strain was isolated from blood cultures.

In the United States (USA), the RB51 vaccine strain replaced the S19 vaccine in 1996, since RB51 was found to be equally immunogenic but less virulent than S19. In 1998, the Centers for Disease Control and Prevention (CDC) published 32 notifications of unintentional inoculation or conjunctival exposure to the RB51 vaccine, occurring in1997 (10). Three of the cases reported inflammation at the inoculation site, and another person described systemic symptoms.

Subsequently, Ashford et al. (2004) published findings from the CDC registry involving reports received from 26 veterinarians accidentally exposed to RB51 during animal vaccination, between 1998 and 2002 (11). 19 of them cited local or systemic symptoms, while 7 reported no adverse events associated with the accidental exposure. Only one of the veterinarians showed a positive culture, which was taken from the cutaneous injection site. Since current standard serologic assays cannot detect antibodies against RB51 infection, and the passive surveillance registry probably underestimates rates of needlestick injuries, the authors concluded that we cannot yet determine whether the RB51vaccine has the potential to cause systemic brucellosis in humans.

In 2005, Avdikou et al. described the results of a local brucellosis surveillance system implemented in a defined region of Northwestern Greece (12). Of a total of 152 newly diagnosed cases recorded during a 2-year study period, 41 (27%) reported contact with the Rev-1 vaccine during its administration.

In Argentina, Wallach et al. (2008) evaluated the pathological consequences of exposure to the vaccine strain *Brucella abortus* S19 in 30 employees from vaccine manufacturing plants, between 1999 and 2006 (13). Fifteen out of 21 laboratory employees with serologically-defined active infection showed clinical manifestations. Blood cultures were performed on nine patients and were negative in all cases. Fever, fatigue, joint stiffness, headache, muscle aches and neuropsycological symptoms were the most frequent findings. Only five of these workers recalled an accidental exposure, indicating that employees from laboratories producing the S19 vaccine are at risk of exposure to *Brucella abortus* by definition, and may become infected by this strain.

Gunes et al. published a serosurveillance study of 46 veterinary staff assigned to a sheep vaccination campaign in Turkey using the Rev.1 strain, in 2013 (14). Ten persons became seropositive (Rose Bengal and Wright tests), but only 2 developed symptoms of infection and were treated with antibiotics. At 6 months, all of them showed negative serology (Wright test). The study suffers from some limitations since serologic tests were not performed prior to the vaccination campaign, and the prevention measures applied were not stated.

Between 2015 and 2016, Proch et al conducted a study aimed to identify risk factors associated with occupational *Brucella* infection in 296 veterinary personnel in India (15). Blood samples were taken from 279 individuals and the Rose Bengal, standard tube agglutination (STAT) and ELISA tests were performed. Previous *Brucella* needlestick injury with the S19 strain was reported in 12 individuals, of whom 5 had a positive serologic test for *Brucella*. After adjusting for other variables, the odds of having a positive serologic test were higher for non-veterinarians, individuals with more seniority and, paradoxically, for those using personal protective equipment (PPE). However, only 29/275 (11%) subjects used PPE, and the appropriateness of its use could not be assesed.

Two cases were subsequently reported that reinforce the danger of occupational exposure to these *Brucella* vaccine strains. The first one occurred in 2017 (16), in a veterinarian in Oregon who developed cough, fever and arthralgia. Following a chest x-ray, the patient was discharged with a doxycycline prescription for pneumonia, but *Brucella* testing was not performed. Four days later, he returned with worsening pneumonia and was hospitalized with doxycycline, rifampicine, ceftriaxone and azithromycin treatment. The Oregon Health Authority Public Health Division (OPHD) was notified of a probable B51 brucellosis case, and treatment was changed to trimethroprim-sufamethoxazole and doxycycline, which cured the patient. However, since blood and sputum cultures were negative for *Brucella spp*, the case could not be bacteriologically confirmed. In 2020, Arapovic reported the first case of Rev.1 human brucellosis in Bosnia and Herzegovina (17); the patient, a farmer, had assisted the veterinarian in vaccinating his sheep, without wearing any PPE. The diagnosis was made by blood culture isolation of the Rev.1 strain, which was identified by molecular methods (multiplex PCR).

Recently, in 2021, Pereira et al. published the results of an online questionnaire carried out on veterinarians registered to administer S19 and RB51 vaccines in Minas Gerais state, Brazil (18). Three hundred and twenty-nine veterinarians were included in the analyses, using stratified random sampling. One hundred and eight (33%) of them cited accidental exposure to S19 or RB51 vaccine strains, 15 (4.6%) reported having brucellosis, and 7 of those 15 considered that the infection was due to accidental exposure to *Brucella* vaccines. Poor knowledge of human brucellosis symptoms and lack of appropriate personal protective equipment (PPE) use were risk factors for unintentional contact with S19 and RB51 vaccine strains.

In 2021, a Spanish study by Vives-Soto et al. analyzed the human serologic response over time in a cohort of vaccine manufacturing workers exposed to the *Brucella melitensis* Rev.1 vaccine strain and, to a much lesser degree, the *Brucella abortus* S19 vaccine (19). Although none of the workers developed symptomatic brucellosis, seropositivity was observed in 47 (41%) of the 115 individuals examined, indicating asymptomatic infection with the vaccine strains, despite strict safety measures. This seropositivity was significantly associated with higher levels of exposure to the vaccine strains. The fact that none of the workers studied developed the disease could be explained by the lower virulence of both strains and / or by a smaller bacterial inoculum due to safety measures. Moreover, the seroconversion of these workers may have actually protected them from contracting brucellosis, as previously described by Buchanan et al. (20).

## DISCUSSION

In this review, we found that *Brucella* vaccines are dangerous to humans and can infect them through aerosol exposure and by mucosal or non-intact skin contact with live attenuated strains. Of the 3 vaccine strains currently used (Rev.1, S19 and RB51), Rev-1 seems to be the most virulent. Prolonged contact with the pathogen, lack of information and instructions provided to the occupational groups exposed and low adherence to personal protective equipment (PPE) in the work environment, appeared to be the main risk factors leading to infection by these vaccine strains. Among veterinarians, vaccine handling was the most reported source of exposure to *Brucella*. On the other hand, despite strict protection measures, a percentage of vaccine manufacturing workers developed positive serology to the vaccine strain, which may have conferred immunity to brucellosis.

The danger of these *Brucella* vaccine strains for use in human inoculation has been studied in various clinical trials. Between 1952 and 1958, Vershilova et al conducted the largest clinical trial on 3 million people engaged in the livestock/meat/food processing industries in the former Soviet Union, using the S19 strain for human vaccination (21). They observed an almost 60% reduction in cases of human brucellosis, with a high safety rate for the vaccine. However, in the Spink et al clinical trial of the *Brucella* vaccines in 1962 (3), 11 of the 16 volunteers receiving the Rev-1 strain developed acute brucellosis, four of them requiring hospitalization; and 4 of the 16 individuals receiving the S19 vaccine reported undesirable sequelae. In 1992, in France, Strady et al carried out a prospective phase IV study with the S19 strain, on 161 professionally exposed human volunteers (22). The authors observed local pain after injection in 45% of subjects and systemic reactions in 5%; however, the clinical efficacy of the vaccine could not be evaluated due to an insufficient number of participants. Lastly, in 1994, Hadjichristodoulou et al conducted a clinical trial on 271 volunteers in Greece; although the S19 vaccine caused some side effects in a quarter of subjects, it was considered safe enough for use on a large scale (23). Nonetheless, there is currently no licensed anti-*Brucella* vaccine for humans. Having said that, at the present time, research is being conducted on developing safe, effective, cross-protecting, exclusively human vaccines due to *Brucella’s* zoonotic potential and possible use in bio-warfare (24).

Some reports emphasize the danger of indirect exposure to *Brucella* vaccine strains; they were not included in this review, since cases were not due to exposure during vaccine manufacturing or administration. In 1998, one such report by the CDC described the case of a stillborn calf, delivered by cesarean. The necropsy revealed that death was due to infection by the RB51 strain, *Brucella abortus* (10). The strain was isolated from placental and fetal lung tissue, as well as from the blood of the calf’s mother and was identified by molecular methods (PCR). The nine persons who participated in the procedures received post-exposure prophylaxis, and none developed brucellosis during the 6-month follow-up period. Later, in 2018, Cossaboom et al reported a case of human brucellosis associated with the consumption of raw (unpasteurized) cow’s milk purchased from a dairy in Paradise, Texas (26). The CDC’s Bacterial Special Pathogens Branch (BSPB) confirmed the isolate as *Brucella abortus* vaccine strain RB51. Recently, in 2021, Sarmiento-Clemente et al. reported the first case in Houston of neurobrucellosis due to the vaccine strain RB51 (26). The patient was an 18-year old Hispanic female who had consumed unpasteurized cheese brought from Mexico 1 month before onset of symptoms. The strain was isolated in blood cultures and identified by molecular methods in the Houston Health Department/Laboratory. The authors also mentioned that the CDC reported 3 confirmed cases in the United States of human infection by RB51 through consumption of raw milk.

Two meta-analyses on the occupational risks of contracting brucellosis have been published, with some references to the risks that *Brucella* vaccine handling poses for humans. One of these, by Xie et al (2018), analyzed 27 papers reviewing adverse effects in animals and humans associated with the three licensed brucellosis vaccines: S19, Rev.1 and RB51 (27). They reported that in animals, adverse effects were mainly found in the immune and reproductive systems, while human adverse effects from occupational exposure to the vaccines usually involved behavioral and neurological systems. The meta-analysis was limited because the majority of the papers included were case reports or case-control studies, and a causal relationship could not be fully established. The other systematic review was published by Pereira et al, in 2020 (28). They addressed the main occupations affected by *Brucella* spp infection, the main risk factors for contracting the disease and the most common forms of exposure to the pathogen, including *Brucella* vaccine strains. Regarding laboratory staff, most of the publications found by them referred to accidents in clinical laboratories but not to manufacturing vaccine exposure. The Pereira et al study was limited by the small number of high-quality papers eligible for meta-analysis and the impossibility of accessing data on the exposed and non-exposed individuals. New evidence has recently been published that sheds light on the risk of infection to humans by these vaccine strains, as shown above in the Results section (18,19).

Our results confirm that there is a clear risk of infection by *Brucella* vaccine strains, although the small number of symptomatic infections recorded compared to the enormous number of doses administered, indicates that most of them are subclinical. Therefore, in order to protect workers from being infected by these vaccine strains, all individuals exposed to Rev.1, S19 or RB51 *Brucella* strains should be considered in a high-risk category, and procedures should be implemented to minimize spills, splashes and aerosols, as well as accidental needlesticks.

Appropriate PPE for vaccination or close exposure to vaccinated animals must include gloves, closed footware, eye protection, a face shield and respiratory protection (6). During the vaccine manufacturing process, strict safety measures in compliance with the Biosafety in Microbiological and Biomedical Laboratories (BMBL) standards are currently in place. They include: a) at least class II Biological Safety Cabinets (BSCs), b) proper personal protective equipment (PPE) and c) use of primary and secondary barriers (29). Following accidental exposure to *Brucella* vaccine strains, symptoms should be monitored and antibiotic post-exposure prophylaxis administered (6). In addition to adequate training for workers, programs monitoring adherence to these standards and quality audits should be implemented.

Notwithstanding the exhaustive protection measures in place, a certain likelihood of exposure persists. Moreover, the applicability of these measures is not always guaranteed, due to human factors such as carelessness, negligence or malingering. Therefore, companies should actively monitor their employees through periodic check-ups that include *Brucella* serology, currently not feasible for RB51, as quality control of the measures applied. In addition, when a worker suffers accidental exposure, a report should be filed, and protocols for medical assessment and post-exposure antibiotic prophylaxis should be activated.

The present review suffers from some limitations. Firstly, there are relatively few published studies on occupational exposure to *Brucella* vaccine strains. In addition, these studies are highly heterogeneous in terms of methodology and diagnostic criteria, as shown in **Table 1**. Moreover, many of them used serology as a diagnostic criterion, which does not distinguish infections caused by vaccine strains from those produced by wild strains. Only two of the studies in our analysis (15, 19) evaluated which protective measures were the most effective for exposed workers, but they were not entirely conclusive due to the difficulty in assessing degree of individual adherence to PPE use.

In conclusion, brucellosis vaccines pose risk of contagion to humans, both from the manufacturing process and their administration to cattle. We recommend extreme measures to avoid or reduce exposure, such as proper use of PPE, employment of primary and secondary barriers and the handling of vaccines in Biological Safety Cabinets. Nonetheless, evidence for the efficacy of these measures is weak, and the incidence of human infection with these vaccines is unclear. Therefore, it would be advisable to carry out observational studies and/or systematic registries using solid diagnostic criteria in populations exposed to *Brucella* vaccine strains. In addition, clinical and serological follow-up programs for exposed workers should be established.

## Data Availability

All data produced in the present study are available upon reasonable request to the authors

## NOTES

### Author Declarations

We confirm that all relevant ethical guidelines were followed. We followed all appropriate research reporting guidelines and uploaded the relevant EQUATOR Network research reporting checklists and other pertinent material as supplementary files, where applicable.

### Availability of all data

All data produced in the present study are available upon reasonable request to the authors.

### Author contributions

Conceptualization and coordination: ERS, JS, JLP; Systematic review and article screening: MVS, APG; Writing – original draft preparation: MVS, APG, JS; Supervision: ERS, JS.

## Acknowledgments

The authors thank Alexandra Salewski Msc for expert English review of the manuscript, and Carlos de Cabo PhD for preparation of the edition and presentation of the work for its publication.

